# How many do we miss? - Evaluation of age at onset and family history as selection criteria for genetic testing in Parkinson’s disease

**DOI:** 10.1101/2025.11.15.25339249

**Authors:** Alexander Balck, Eva-Juliane Vollstedt, Ana Westenberger, Lara M. Lange, Joanne Trinh, Meike Kasten, Katja Lohmann, Norbert Brüggemann, Claudia Trenkwalder, Brit Mollenhauer, Roy Alcalay, Kamalini Ghosh Galvelis, James C. Beck, Peter Bauer, the Global Parkinson’s Genetics Program (GP2), the ROPAD Study Group, the PDGENEration Study, Christine Klein, Inke R. König

## Abstract

**Importance:** Current recommendations for genetic testing in Parkinson’s disease (PD) prioritize groups of patients based on age at onset (AAO) and family history (FH). The increasing importance of identifying genetic PD for personalized counseling and potential gene-specific therapies calls for a data-driven evaluation of these recommendations.

**Objective:** To estimate the diagnostic accuracy, specifically the sensitivity, specificity, and positive predictive value (PPV), of genetic testing in PD based on AAO and FH.

**Design, Setting, and Participants:** We analyzed data from six cohorts within four independent datasets: ROPAD, PD GENEration, the MDSGene database, the Global Parkinson’s Genetics Program (GP2), and two German observational studies. These datasets included 25,063 PD participants, of whom 6,295 carried pathogenic or likely pathogenic variants. ROPAD and PD GENEration, both prospective genetic screening studies, served as representative real-world cohorts.

**Main Outcome(s) and Measure(s):** For each gene, we quantified the proportion of carriers by age bracket and familial vs. sporadic status. Receiver operating characteristic (ROC) curves, and area under the curve (AUC) values with 95% confidence intervals (CIs) were calculated for AAO and FH. PPVs were computed based on sensitivity, specificity, and prevalence.

**Results:** An AAO threshold of ≤50 identified 32% (ROPAD), 23% (PD GENEration), 63% (MDSGene), and 30% (GP2) of genetic cases. ROC analyses based on AAO alone yielded AUCs of 0.59 (CI: 0.57-0.60, ROPAD), 0.58 (CI: 0.56-0.60, PD GENEration), 0.78 (CI: 0.75-0.80, MDSGene), and 0.54 (CI: 0.52-0.56, GP2), respectively. Combining AAO with FH increased AUCs to 0.60 (CI: 0.59-0.62), 0.60 (CI: 0.58-0.62), 0.83 (CI: 0.81-0.85), and 0.58 (CI: 0.56-0.60). FH improved AUCs for dominant genes such as *LRRK2* (e.g., 0.52 to 0.61 in ROPAD) but had minimal or no effect for recessive genes (e.g., *PRKN/PINK1*: 0.90 to 0.90 in ROPAD).

**Conclusions and Relevance:** Current selection criteria (AAO ≤50) identify only a minority (23–32%) of variant carriers. Most carriers (68–77%) present with a later AAO and remain undetected. While AAO is moderately predictive in some cohorts, it insufficiently captures late-onset genetic forms, particularly *LRRK2*-PD and *GBA1*-PD. The limited incremental value of FH challenges its use as a selection criterion. These findings support revised, data-driven testing strategies to improve detection of genetic PD across all age groups.

**Key Points:** 

**Question:** What are the diagnostic accuracy, specifically the sensitivity, specificity, and positive predictive value, of the current age at onset and family history-based selection criteria for genetic testing to identify major gene pathogenic variant carriers among patients with Parkinson’s disease?

**Findings:** Performing genetic testing in Parkinson’s disease patients with an early age at onset ≤50 years results in a positive genetic finding in one of four patients; however, this criterion captures only 23–32% of known variant carriers in population-based datasets, underscoring its limited sensitivity and leading to substantial underdiagnosis of genetic Parkinson’s disease. The additional discriminatory value of family history is marginal.

**Meaning:** Revised recommendations for broader genetic testing in Parkinson’s disease are warranted.

## Introduction

Although genetic factors underlying Parkinson’s disease (PD) have been known for almost 30 years and affect approximately 15% of all PD patients^1,2^, there are no widely accepted data-based recommendations as to which PD patients should undergo genetic testing.^3^ Existing guidance from professional societies, such as the Movement Disorder Society (MDS) and the European Federation of Neurological Societies (EFNS), recommends genetic testing based on age at onset (AAO) and family history (FH), often specifying it for individual genes or modes of inheritance.^3,4^ Some groups suggest genetic testing in every PD patient with an AAO ≤50 years, regardless of FH, ethnicity, and presumed inheritance pattern,^3,5^ and others suggest incorporating genetic testing into routine PD patient care.^6^ These approaches are based on expert opinion, and their diagnostic accuracy, specifically, sensitivity, specificity, and positive predictive value (PPV), have not been assessed.^3^ AAO varies substantially across genetic forms of PD, limiting its utility as a uniform selection criterion for genetic testing.^7,8^

Although individual penetrance and expressivity of variants in monogenic PD genes (such as *PRKN, PINK1, PARK7, LRRK2, SNCA, and VPS35*) and in *GBA1* – a relatively common genetic risk factor for PD – cannot be predicted with certainty, genotype information remains clinically meaningful for risk assessment, counselling, and prognostication.^9^ It may also inform clinical decisions, as it is under debate whether *GBA1* variant carriers face a higher risk of cognitive decline following stimulation.^9–12^ These insights will become even more consequential as the first gene-specific therapies are developed, especially for *LRRK2*- and *GBA1*-related PD.

Given the growing clinical relevance of identifying genetic factors underlying PD on an individual level, and as genetic testing becomes more comprehensive and more widely affordable, a data-driven assessment of testing guidelines is warranted. Notably, most of the available evidence, including all data from the ROPAD and PD GENEration studies, was generated from clinical-grade genetic testing with return of results to participants, underscoring the value and feasibility of such approaches in clinical practice. In this study, we estimate the diagnostic accuracy of utilizing AAO and FH as selection criteria for PD genetic testing, focusing on sensitivity, specificity, and PPV.

## Methods

We used four independent datasets, derived from six distinct sources, to evaluate the sensitivity, specificity, and PPV of AAO and FH as selection criteria for genetic testing. These included two prospective clinical-genetic screening studies (ROPAD and PD GENEration), one curated literature-based dataset (MDSGene), two genetically characterized observational cohorts of idiopathic PD patients (EPIPARK and DeNoPa), and a globally aggregated genotyping and sequencing resource including both monogenic and idiopathic PD (IPD) patients (Global Parkinson’s Genetics Program [GP2]). Information on study design, participant selection, and data collection is provided in the Supplement (**eMethods 1-4**) and the original publications.^1,2,13–16^

ROPAD and PD GENEration are prospective genetic screening studies including AAO, FH, and genetic data. Both served as “real-world” data sources, enabling estimation of genetic PD’s prevalence and subsequent PPV estimates. Neither study was specifically enriched for early onset or familial patients, although patients with a FH or lower AAO were more likely to be recruited (**eMethods 1-2**).^1,2,13^

The MDSGene database collects genetic and phenotypic information on PD participants carrying pathogenic variants reported in the English-language literature^17–19^. Since it only contains participants with genetic forms of PD, we complemented it with IPD data from EPIPARK^14^ and DeNoPa^15^. PD gene panel analyses confirmed that none of these IPD participants carried pathogenic variants in PD-causing genes (**eMethods 3**). Together, these datasets were analyzed as the combined ‘MDSGene dataset’.

GP2 provided the fourth dataset (**eMethods 4**), excluding individuals from the aforementioned studies to avoid duplicates. Duplicates with MDSGene cannot be excluded entirely. Participants in GP2 cohorts may have been “pre-screened” for some cohorts, reducing the rate of pathogenic variants.

Participants with missing data for AAO or with pathogenic variants in more than one gene, benign variants, or variants of unknown significance (**eFigure 1**) were excluded.

AAO was defined as the manifestation of the first cardinal sign(s) of PD and was determined based on the patient’s history. For PD GENEration, FH was considered positive if a first-degree family member had PD. The other datasets considered FH positive if PD was reported in any family member.

Receiver operating characteristic (ROC) curves based on AAO were used to differentiate between genetic PD and IPD, and the areas under the curve (AUCs) were estimated with 95% confidence intervals (CI)^20^. Logistic-regression models combining AAO and FH were applied to compute AUCs for composite predictors, with DeLong tests for pairwise comparisons.

Precision–recall curves were generated to address class imbalance. Sensitivity and PPV were calculated for various AAO thresholds and ranges; PPV estimation was limited to ROPAD and PD GENEration, where mutation frequencies were known. Gene-specific analyses were performed to differentiate the most frequent forms of autosomal recessive PD (*PRKN*-PD, *PINK1*-PD, *PARK7-PD*), the most frequent form of autosomal dominant PD (*LRRK2*-PD), and the most common genetic risk factor for PD (*GBA1*-PD) from IPD participants based on AAO and FH.

## Results

The ROPAD dataset consisted of 12,209 individuals (*PRKN*: n=118, *PINK1*: n=9, *DJ-1*: n=4, *LRRK2*: n=368, *SNCA*: n=25, *VPS35*: n=2, *GBA1: n=1,306*, and IPD: n=10,377, **eFigure 1**). Monogenic and *GBA1*-related PD represented 15.0% (n=1,832) of the dataset. FH data for IPD were available for 10,045 participants and were positive in 26.1% of the cohort (n=2,623/10,045). For monogenic and *GBA1*-related PD, data on FH were available for 1,768 participants and were positive in 36.9% (n=645/1,768, **Table 1**).

**Table1.** Number of included individuals, age at onset (AAO), and positive family history (FH) per dataset and gene. Percentages of individual genes refer to the total number of individuals, including IPD. Percentages for positive family history refer to the patients with positive family history divided by the patients with information on FH per group. AAO values are the median with the respective IQR. *IPD patient data in the MDSGene dataset originate from EPIPARK and DeNoPa (see methods).

|  | ROPAD dataset |  |  | PD GENERation dataset |  |  | MDSGene dataset |  |  | GP2 dataset |  |  |
| --- | --- | --- | --- | --- | --- | --- | --- | --- | --- | --- | --- | --- |
|  | n (%) | AAO (IQR) | FH pos. (%) | n (%) | AAO (IQR) | FH pos. (%) | n (%) | AAO (IQR) | FH pos. (%) | n (%) | AAO (IQR) | FH pos. (%) |
| **GBA1** | 1,306 (10.7) | 56 (48 – 63) | 404/1,256 (32.2) | 620 (7.9) | 59 (52 – 66) | 131/592 (22.1) | 563 (22.2) | 51 (43 – 60) | 133/285 (46.6) | 802 (17.3) | 57 (48 – 66) | 234/711 (32.9) |
| **LRRK2** | 368 (3.0) | 58 (50 – 65) | 161/355 (45.4) | 194 (2.5) | 63 (57 – 69) | 76/186 (40.1) | 800 (31.5) | 56 (46 – 63) | 359/587 (61.1) | 208 (4.5) | 57 (49 – 66) | 110/192 (57.3) |
| **SNCA** | 25 (2.0) | 50 (40 – 59) | 18/27 (66.7) | 9 (0.1) | 47 (41 – 54) | 7/11 (64.6) | 80 (3.2) | 48 (40 – 59) | 96/118 (80.5) | 1 (<0.1) | 52.5 (44 – 64) | 2/2 (100) |
| **VPS35** | 2 (<0.1) |  |  | 4 (0.1) |  |  | 48 (1.9) |  |  | 3 (0.1) |  |  |
| **PRKN** | 118 (1.0) | 35 (29 – 43) | 62/130 (47.7) | 56 (0.7) | 35.5 (27 – 48) | 28/61 (45.9) | 882 (34.8) | 30 (23 – 38) | 456/774 (58.9) | 8 (0.2) | 38 (29 – 46) | 16/22 (72.7) |
| **PINK1** | 9 (0.1) |  |  | 4 (0.1) |  |  | 127 (5.0) |  |  | 15 (0.3) |  |  |
| **PARK7** | 4 (<0.1) |  |  | 2 (<0.1) |  |  | 37 (1.5) |  |  | - |  |  |
| **All genetic (without GBA1)** | 526 (4.3) | 53 (42 – 63) | 241/512 (47.1) | 269 (3.4) | 60 (48 – 67) | 111/258 (43.0) | 1974 (68.3) | 40 (29 – 55) | 911/1479 (61.6) | 235 (5.1) | 56 (46 – 65) | 128/216 (59.2) |
| **All genetic** | 1,832 (15.0) | 55 (47 – 63) | 645/1,768 (36.9) | 889 (11.3) | 59 (51 – 67) | 242/850 (28.5) | 2,537 (87.8) | 43 (32 – 57) | 1,044/1,764 (59.2) | 1,037 (22.4) | 57 (48 – 66) | 362/927 (39.1) |
| **IPD** | 10,377 (85.0) | 59 (50 – 67) | 2,623/10,045 (26.1) | 7,003 (88.7) | 63 (55 – 69) | 1,282/6,741 (19.0) | 351* (12.2) | 62 (52 – 69) | 75/329 (22.8) | 3,602 (77.6) | 60 (50 – 67) | 904/3,225 (28.0) |

The PD GENEration dataset consisted of 7,892 individuals (*PRKN* n=56, *PINK1* n=4, *PARK7* n=2, *LRRK2* n=194, *SNCA* n=9, *VPS35* n=4, *GBA1* n=620, IPD n=7,003, **eFigure 1**). Monogenic and *GBA1*-related PD represented 11.3% (n=889) of the dataset. Data on FH were available for 7,591 participants (96.2%), and first-degree relative FH was positive in 19.0% of IPD cases (n=1,282/6,741) and 28.5% of the genetic PD patients (n=242/850, **Table 1**).

For the MDSGene dataset, information on AAO was available for 2,537 index patients with a genetic cause of PD (*PRKN*: n*=882, PINK1*: n=127*, PARK7*: n=37, *LRRK2*: n=800, *SNCA*: n=80, and *VPS35*: n=48, *GBA1:* n=563). Data on FH were available for 1,764 participants and positive in 59.2% of them (n=1,044/1,764). AAO data of 351 IPD patients were obtained from the EPIPARK and DeNoPa cohorts, representing 12.2% of the MDSGene dataset (n=351/2,888). Data on FH were available in 329 cases and positive in 22.8% (n=75/329, **Table 1**).

In the GP2 dataset, AAO data were available for 3,602 IPD participants, 3,225 of whom also had FH data. FH was positive for 28.0% (n=904/3,225). AAO data were available for 1,037 monogenic and *GBA1*-related PD participants (*PRKN*: n*=*8*, PINK1*: n=15*, PARK7*: n=0, *LRRK2*: n=208, *SNCA*: n=1, *VPS35*: n=3, *GBA1*: n=802), of which 927 also had FH data. FH was positive for 39.1% (n=362/927, **Table 1**).

Compared to IPD, the median AAO for the genetic PD participants was lower in all four datasets (ROPAD dataset: 59 years [interquartile range: 50-67 years] *vs.* 55 years [interquartile range: 47-63 years]; PD GENEration dataset: 63 years [interquartile range: 55-69 years] *vs.* 59 [interquartile range: 51-67 years]; MDSGene dataset: 62 years [interquartile range: 52-69 years] *vs.* 43 years [interquartile range: 32-57 years]; GP2 dataset: 60 years [interquartile range: 50-67 years] *vs.* 57 years [interquartile range: 48-66 years], **Table1**).

Based on AAO, a differentiation of PD patients into ‘any’ genetic *vs.* IPD patients was possible (AUC: 0.59 (CI: 0.57-0.60, ROPAD dataset); 0.58 (CI: 0.56-0.60, PD GENEration dataset); 0.78 (CI: 0.75-0.80, MDSGene dataset); 0.54 (CI: 0.52-0.56, GP2 dataset), **Table 2**, **Figure 1**). A regression model classifying carrier status from AAO and FH generated a marginally larger AUC for the “real-world” studies (0.60 [CI: 0.59-0.62] ROPAD dataset, 0.60 [CI: 0.58-0.62] PD GENEration dataset), and a considerably larger AUC for the MDSGene and GP2 dataset (0.83 [CI. 0.81-0.85] and 0.58 [CI: 0.56-0.60], respectively; **Table 2**, **Figure 1**).

**Figure 1:**
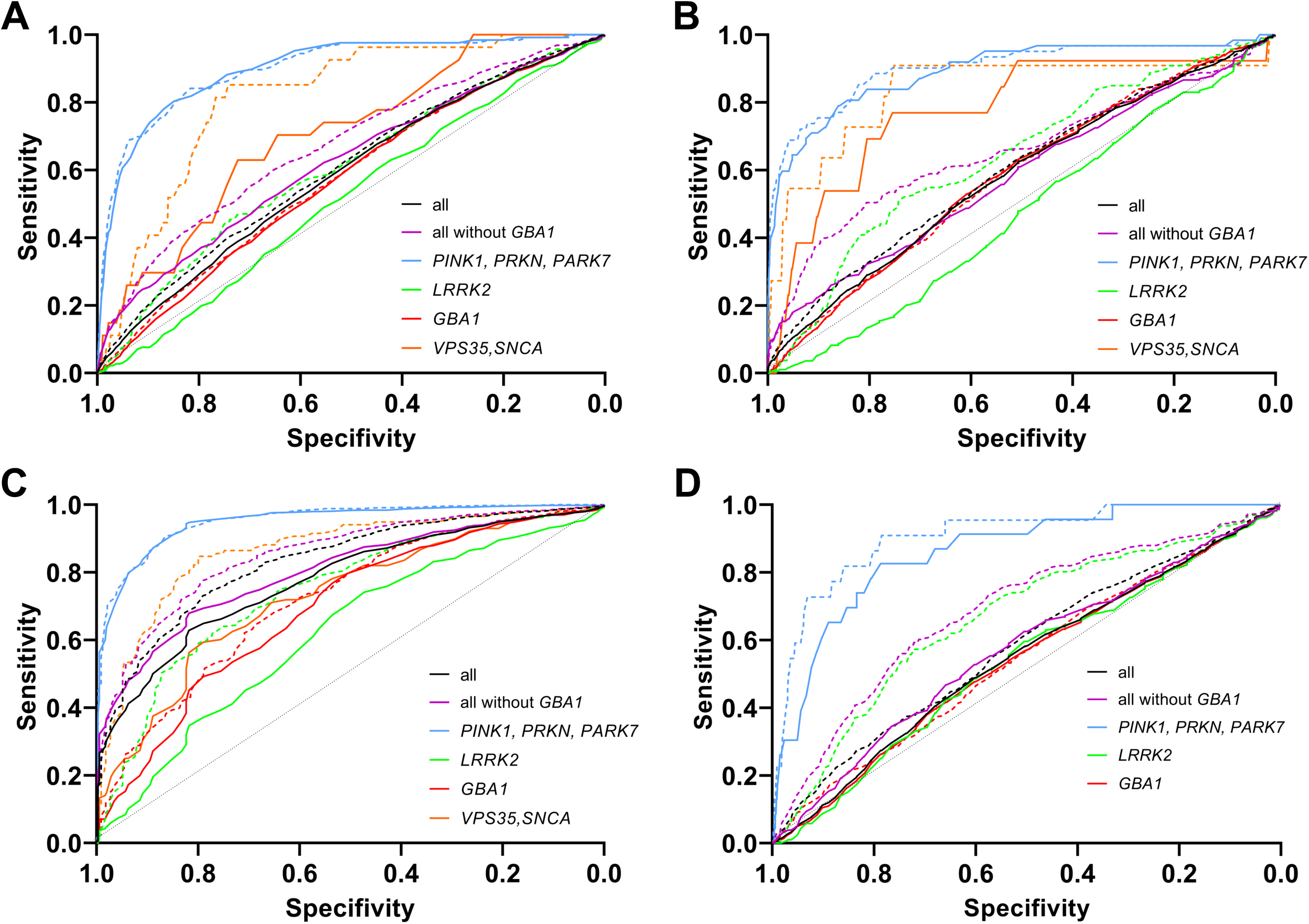
ROC curves of genetic testing specificity and sensitivity based on age at onset (AAO) and family history (FH). A: ROPAD dataset. B: PD GENEration dataset. C: MDS Gene dataset. D. GP2 dataset. Black solid curve: Differentiation of all genetic PD from IPD patients based on AAO. Black dashed curve: Differentiation of all genetic PD patients from IPD patients based on AAO and FH. Purple solid curve: Differentiation of all genetic PD without *GBA1* patients from IPD patients based on AAO. Purple dashed curve: Differentiation of all genetic PD patients without *GBA1* patients from IPD patients based on AAO and FH. Blue solid curve: Differentiation of *PINK1, PRKN, and PARK7* carriers from IPD patients based on AAO. Blue dashed curve: Differentiation of *PINK1, PRKN, and PARK7* carriers from IPD patients based on AAO and FH. Green solid curve: Differentiation of *LRRK2* carriers from IPD patients based on AAO. Green dashed curve: Differentiation of *LRRK2* carriers from IPD patients based on AAO and FH. Red solid curve: Differentiation of *GBA1* carriers from IPD patients based on AAO. Red dashed curve: Differentiation of *GBA1* carriers from IPD patients based on AAO and FH.

**Table 2.**
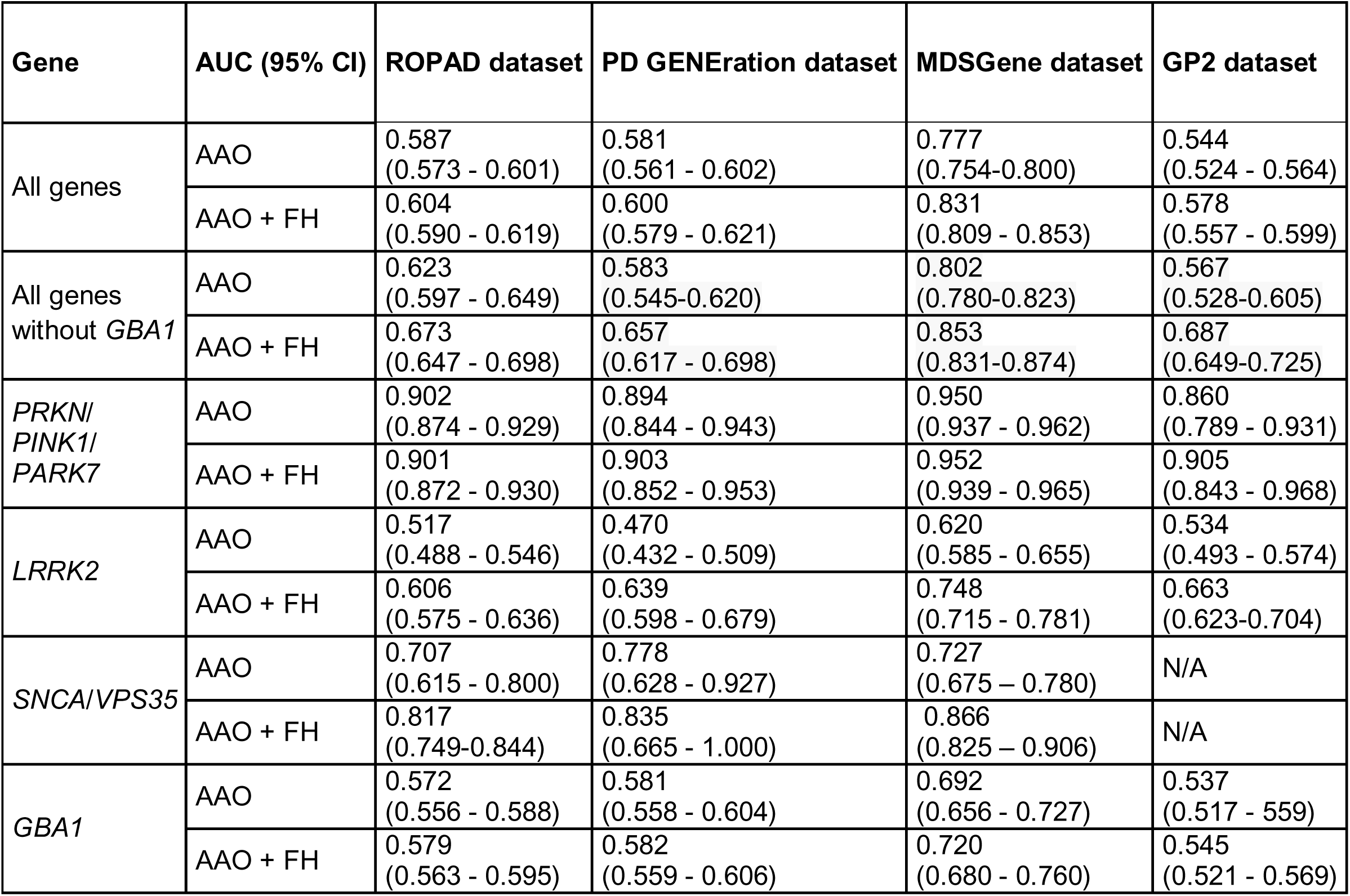
Area Under the Curve (AUC) estimates for genetic Parkinson’s Disease prediction based on age at onset and family history across four cohorts. Differentiation into genetic and IPD is based on age at onset (AAO), and AAO with family history (FH) simultaneously. N/A = no calculations were performed due to the small n

In both real-world datasets, adding FH to AAO significantly increased the AUC (ROPAD: ΔAUC=0.015, 95%□CI=0.006 to 0.023, □DeLong p<0.001; PD GENEration: ΔAUC=0.019, 95% CI=0.006 to 0.032, DeLong□p=0.005). Precision–recall AUCs indicated moderate performance under class imbalance (0.20 and 0.17, respectively; **eTable2**). In the MDSGene dataset, the AUC increase from AAO to AAO□+□FH was also statistically significant (ΔAUC=0.043, 95% CI□=0.027 to 0.058, DeLong p<0.001), and the precision-recall AUC was 0.96, reflecting high performance given the high prevalence of carriers in this curated dataset (**eTable2**). In GP2, a significant AUC increase was also observed (ΔAUC=0.026, 95% CI =0.008 to 0.044, DeLong p=0.005), with a corresponding precision-recall AUC of 0.24, slightly above the carrier prevalence of 22% (**eTable2**).

When focusing on genes linked to early-onset PD, i.e., *PRKN, PINK1, PARK7*, the AUC of differentiating genetic PD from IPD based on AAO was greater (ROPAD dataset: 0.90 [CI: 0.87-0.93]; PD GENEration dataset: 0.89 [CI: 0.84-0.94]; MDSGene dataset: 0.95 [CI: 0.94-0.96], and GP2 dataset: 0.86 [CI: 0.79-0.93]). A regression model based on AAO and FH generated very similar AUCs (0.90 [CI: 0.87-0.93, ROPAD dataset]; 0.90 [CI: 0.85-0.95, PD GENEration dataset]; 0.95 [CI: 0.94-0.97, MDSGene dataset]; 0.91 [CI: 0.84-0.97, GP2 dataset]; **Table 2**, **Figure 1**).

In contrast, when comparing IPD with *LRRK2*-PD, the AUC amounted to 0.52 (CI: 0.49-0.55, ROPAD dataset) when considering AAO (PD GENEration dataset: 0.47, CI: 0.43-0.51; MDSGene dataset: 0.62, CI: 0.59-0.66; GP2 dataset: 0.53, CI: 0.49-0.57). A regression model incorporating both AAO and FH resulted in larger AUCs (ROPAD dataset: 0.61, CI: 0.58-0.64; PD GENEration dataset: 0.64, CI: 0.60-0.68, MDSGene dataset: 0.75, CI: 0.72-0.78, GP2 dataset: 0.66, CI: 0.62-0.70; **Table 1, Figure 1**).

The sensitivity, specificity, and PPV of genetic testing at various AAO thresholds are displayed in **Table 3** and **Figure 2**. The sensitivity increased with the AAO threshold and was generally higher for *PRKN, PINK1*, and *PARK7* than for other genes (*VPS35*, *SNCA,* and *GBA1*). At the same time, *LRRK2* had the lowest sensitivity in all samples (**Table 3**, **Figure 2**). As expected, PPV was highest for genes with greater prevalence and declined with increasing age. The difference in PPV of testing with the currently used threshold (AAO ≤50, PPV for all genes in the ROPAD dataset: 0.20; PD GENEration dataset: 0.17) compared to a higher age threshold (AAO ≤60, PPV for all genes in ROPAD dataset: 0.18; PD GENEration dataset: 0.15) was around 2% (**Table 3**, **Figure 2**). PPV estimation across AAO ranges showed substantial values even in older-onset groups: PD GENEration, PPV = 0.10 for AAO 60–70 years and 0.08 for 70–80 years (**eTable 1**). Corresponding performance metrics (AUC differences, DeLong tests, and precision-recall curves) for each genetic subgroup are reported in the Supplement (**eTable 2**).

**Figure 2:**
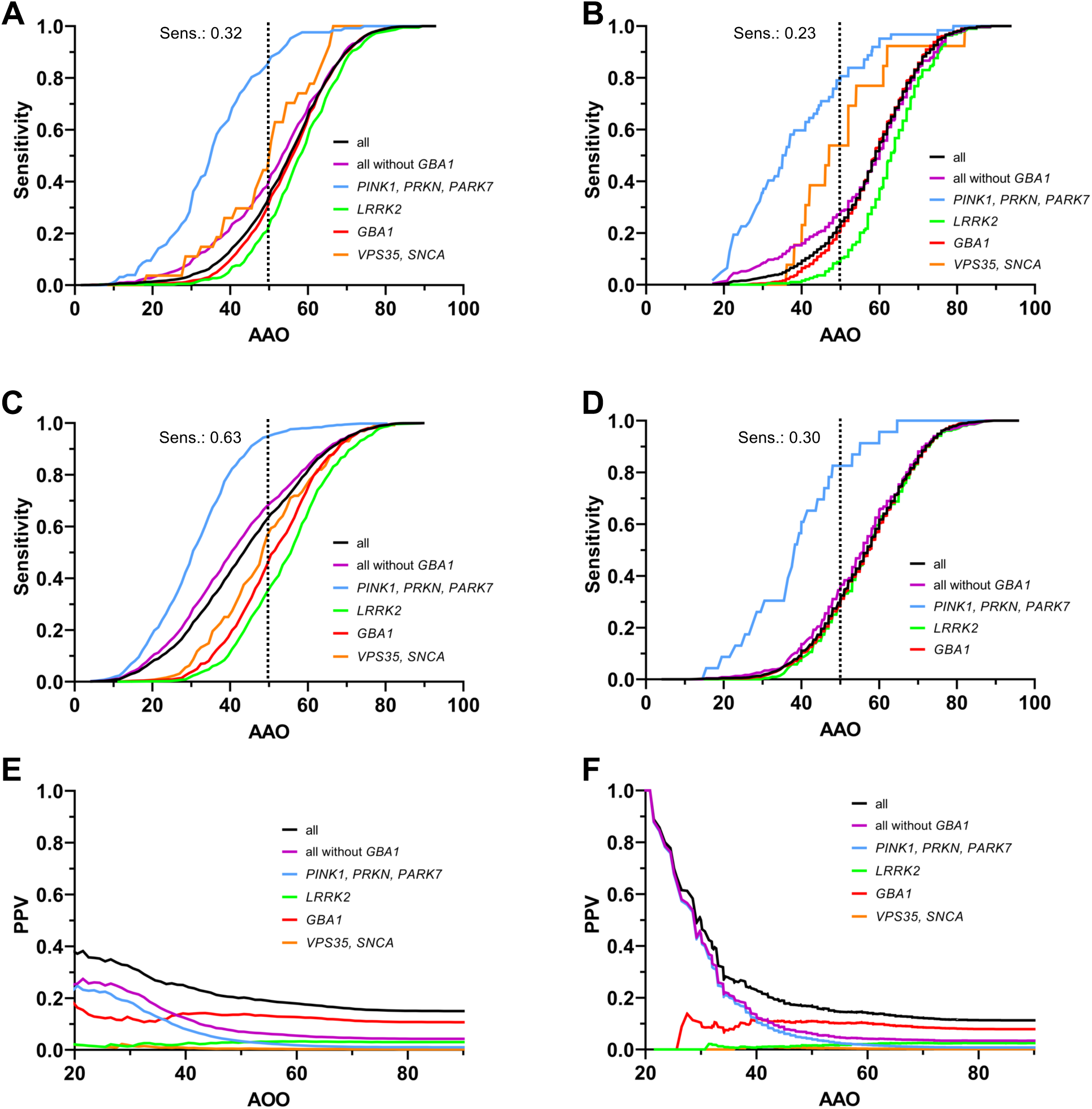
Sensitivity and positive predictive value (PPV) of genetic testing at different age at onset (AAO) thresholds. A: ROPAD dataset. B: PD GENEration dataset. C: MDS Gene dataset. D. GP2 dataset. E: PPV in the ROPAD dataset. F: PPV in the PD GENEration dataset. Panels A-D show the sensitivity of differentiating genetic PD from idiopathic PD (IPD) as a function of the AAO. The dotted black line indicates the sensitivity at the commonly used threshold of AAO ≤50 years. Panels E–F display the PPV as a function of AAO. Black curve: all genetic PD; Blue curve: *PINK1, PRKN, and PARK7* carriers; Green curve: *LRRK2* carriers; Red curve: *GBA1* carriers. Note that the PPV curves reflect the probability of a positive genetic test result at a given AAO threshold, stratified by gene group.

**Table 3:**
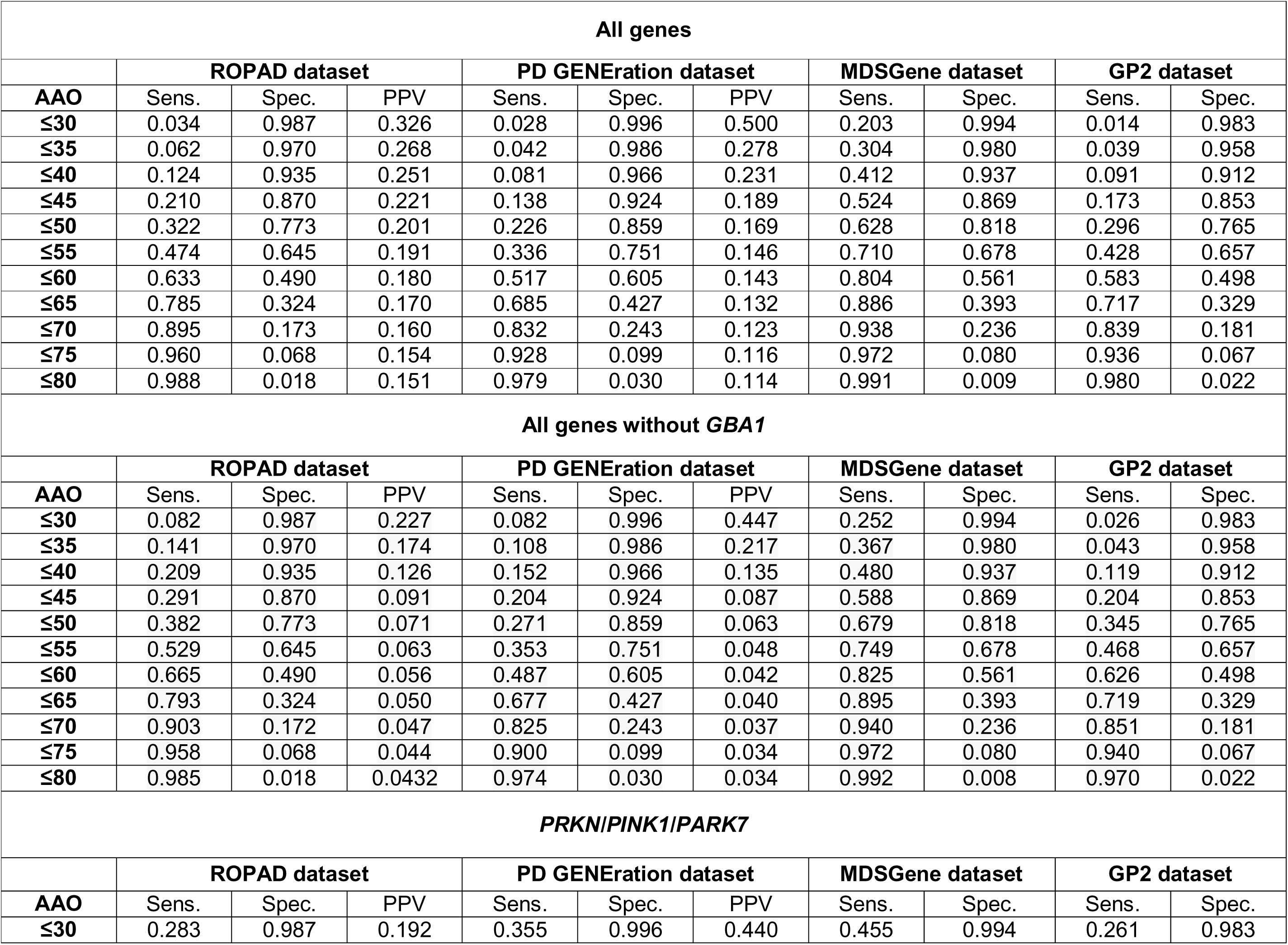
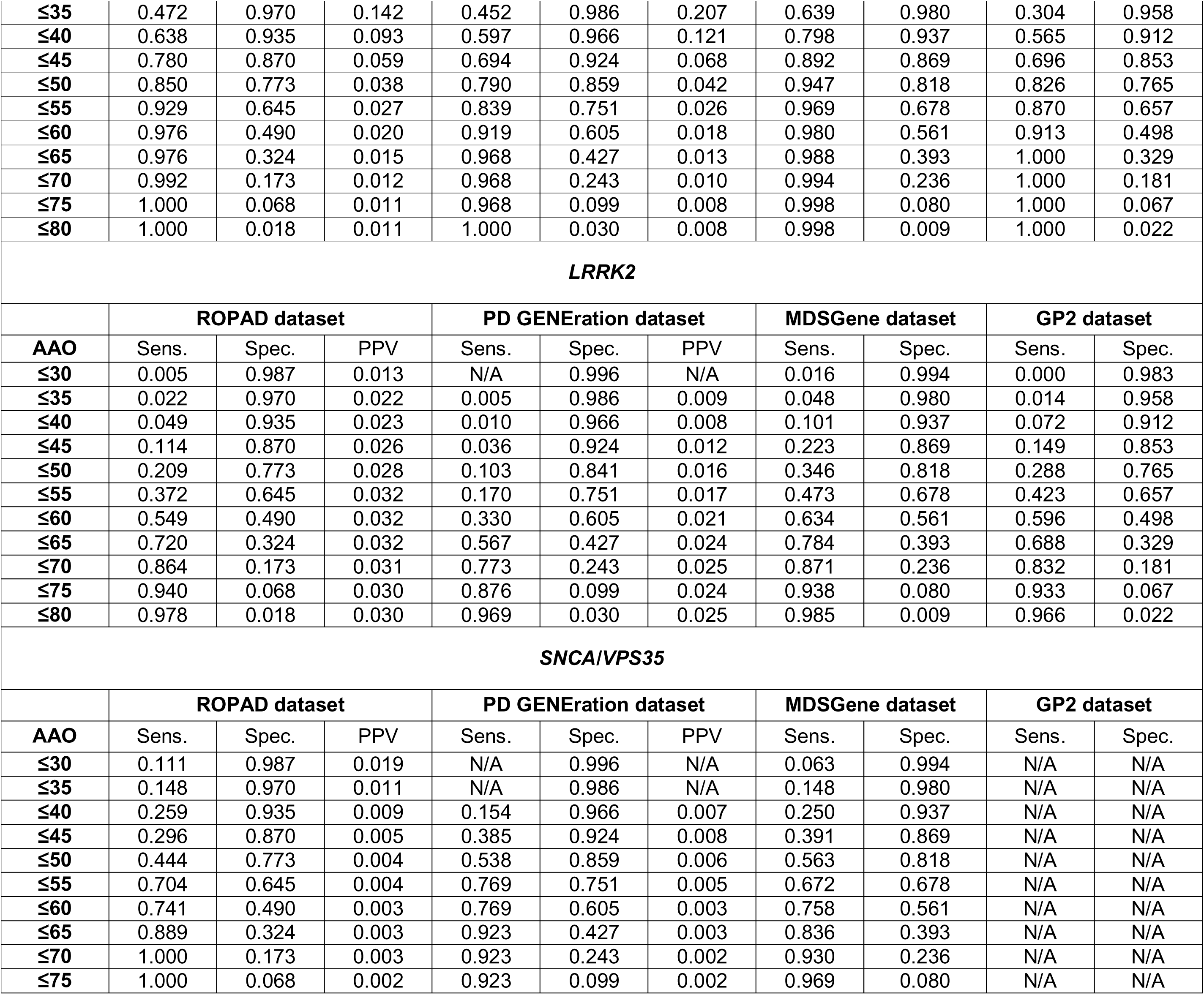
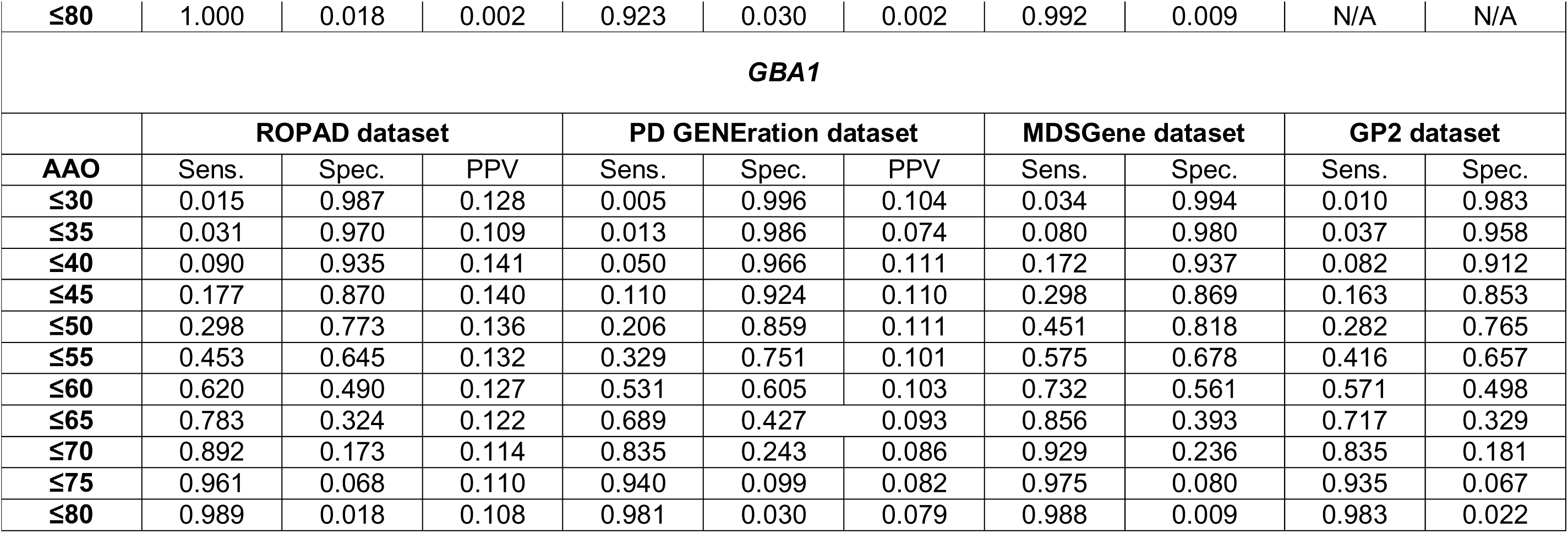
Sensitivity and specificity of genetic testing at different age at onset (AAO) thresholds for all four datasets. Sens. = Sensitivity; Spec. = Specificity; PPV = positive predictive value; N/A = no calculations were performed due to the small n

| All genes |  |  |  |  |  |  |  |  |  |  |
| --- | --- | --- | --- | --- | --- | --- | --- | --- | --- | --- |
|  | ROPAD dataset |  |  | PD GENERation dataset |  |  | MDSGene dataset |  | GP2 dataset |  |
| AAO | Sens. | Spec. | PPV | Sens. | Spec. | PPV | Sens. | Spec. | Sens. | Spec. |
| ≤30 | 0.034 | 0.987 | 0.326 | 0.028 | 0.996 | 0.500 | 0.203 | 0.994 | 0.014 | 0.983 |
| ≤35 | 0.062 | 0.970 | 0.268 | 0.042 | 0.986 | 0.278 | 0.304 | 0.980 | 0.039 | 0.958 |
| ≤40 | 0.124 | 0.935 | 0.251 | 0.081 | 0.966 | 0.231 | 0.412 | 0.937 | 0.091 | 0.912 |
| ≤45 | 0.210 | 0.870 | 0.221 | 0.138 | 0.924 | 0.189 | 0.524 | 0.869 | 0.173 | 0.853 |
| ≤50 | 0.322 | 0.773 | 0.201 | 0.226 | 0.859 | 0.169 | 0.628 | 0.818 | 0.296 | 0.765 |
| ≤55 | 0.474 | 0.645 | 0.191 | 0.336 | 0.751 | 0.146 | 0.710 | 0.678 | 0.428 | 0.657 |
| ≤60 | 0.633 | 0.490 | 0.180 | 0.517 | 0.605 | 0.143 | 0.804 | 0.561 | 0.583 | 0.498 |
| ≤65 | 0.785 | 0.324 | 0.170 | 0.685 | 0.427 | 0.132 | 0.886 | 0.393 | 0.717 | 0.329 |
| ≤70 | 0.895 | 0.173 | 0.160 | 0.832 | 0.243 | 0.123 | 0.938 | 0.236 | 0.839 | 0.181 |
| ≤75 | 0.960 | 0.068 | 0.154 | 0.928 | 0.099 | 0.116 | 0.972 | 0.080 | 0.936 | 0.067 |
| ≤80 | 0.988 | 0.018 | 0.151 | 0.979 | 0.030 | 0.114 | 0.991 | 0.009 | 0.980 | 0.022 |
| All genes without *GBA1* |  |  |  |  |  |  |  |  |  |  |
|  | ROPAD dataset |  |  | PD GENERation dataset |  |  | MDSGene dataset |  | GP2 dataset |  |
| AAO | Sens. | Spec. | PPV | Sens. | Spec. | PPV | Sens. | Spec. | Sens. | Spec. |
| ≤30 | 0.082 | 0.987 | 0.227 | 0.082 | 0.996 | 0.447 | 0.252 | 0.994 | 0.026 | 0.983 |
| ≤35 | 0.141 | 0.970 | 0.174 | 0.108 | 0.986 | 0.217 | 0.367 | 0.980 | 0.043 | 0.958 |
| ≤40 | 0.209 | 0.935 | 0.126 | 0.152 | 0.966 | 0.135 | 0.480 | 0.937 | 0.119 | 0.912 |
| ≤45 | 0.291 | 0.870 | 0.091 | 0.204 | 0.924 | 0.087 | 0.588 | 0.869 | 0.204 | 0.853 |
| ≤50 | 0.382 | 0.773 | 0.071 | 0.271 | 0.859 | 0.063 | 0.679 | 0.818 | 0.345 | 0.765 |
| ≤55 | 0.529 | 0.645 | 0.063 | 0.353 | 0.751 | 0.048 | 0.749 | 0.678 | 0.468 | 0.657 |
| ≤60 | 0.665 | 0.490 | 0.056 | 0.487 | 0.605 | 0.042 | 0.825 | 0.561 | 0.626 | 0.498 |
| ≤65 | 0.793 | 0.324 | 0.050 | 0.677 | 0.427 | 0.040 | 0.895 | 0.393 | 0.719 | 0.329 |
| ≤70 | 0.903 | 0.172 | 0.047 | 0.825 | 0.243 | 0.037 | 0.940 | 0.236 | 0.851 | 0.181 |
| ≤75 | 0.958 | 0.068 | 0.044 | 0.900 | 0.099 | 0.034 | 0.972 | 0.080 | 0.940 | 0.067 |
| ≤80 | 0.985 | 0.018 | 0.0432 | 0.974 | 0.030 | 0.034 | 0.992 | 0.008 | 0.970 | 0.022 |
| *PRKN/PINK1/PARK7* |  |  |  |  |  |  |  |  |  |  |
|  | ROPAD dataset |  |  | PD GENERation dataset |  |  | MDSGene dataset |  | GP2 dataset |  |
| AAO | Sens. | Spec. | PPV | Sens. | Spec. | PPV | Sens. | Spec. | Sens. | Spec. |
| ≤30 | 0.283 | 0.987 | 0.192 | 0.355 | 0.996 | 0.440 | 0.455 | 0.994 | 0.261 | 0.983 |
| ≤35 | 0.472 | 0.970 | 0.142 | 0.452 | 0.986 | 0.207 | 0.639 | 0.980 | 0.304 | 0.958 |
| ≤40 | 0.638 | 0.935 | 0.093 | 0.597 | 0.966 | 0.121 | 0.798 | 0.937 | 0.565 | 0.912 |
| ≤45 | 0.780 | 0.870 | 0.059 | 0.694 | 0.924 | 0.068 | 0.892 | 0.869 | 0.696 | 0.853 |
| ≤50 | 0.850 | 0.773 | 0.038 | 0.790 | 0.859 | 0.042 | 0.947 | 0.818 | 0.826 | 0.765 |
| ≤55 | 0.929 | 0.645 | 0.027 | 0.839 | 0.751 | 0.026 | 0.969 | 0.678 | 0.870 | 0.657 |
| ≤60 | 0.976 | 0.490 | 0.020 | 0.919 | 0.605 | 0.018 | 0.980 | 0.561 | 0.913 | 0.498 |
| ≤65 | 0.976 | 0.324 | 0.015 | 0.968 | 0.427 | 0.013 | 0.988 | 0.393 | 1.000 | 0.329 |
| ≤70 | 0.992 | 0.173 | 0.012 | 0.968 | 0.243 | 0.010 | 0.994 | 0.236 | 1.000 | 0.181 |
| ≤75 | 1.000 | 0.068 | 0.011 | 0.968 | 0.099 | 0.008 | 0.998 | 0.080 | 1.000 | 0.067 |
| ≤80 | 1.000 | 0.018 | 0.011 | 1.000 | 0.030 | 0.008 | 0.998 | 0.009 | 1.000 | 0.022 |

**LRRK2**
|  | ROPAD dataset |  |  | PD GENERation dataset |  |  | MDSGene dataset |  | GP2 dataset |  |
| --- | --- | --- | --- | --- | --- | --- | --- | --- | --- | --- |
| AAO | Sens. | Spec. | PPV | Sens. | Spec. | PPV | Sens. | Spec. | Sens. | Spec. |
| ≤30 | 0.005 | 0.987 | 0.013 | N/A | 0.996 | N/A | 0.016 | 0.994 | 0.000 | 0.983 |
| ≤35 | 0.022 | 0.970 | 0.022 | 0.005 | 0.986 | 0.009 | 0.048 | 0.980 | 0.014 | 0.958 |
| ≤40 | 0.049 | 0.935 | 0.023 | 0.010 | 0.966 | 0.008 | 0.101 | 0.937 | 0.072 | 0.912 |
| ≤45 | 0.114 | 0.870 | 0.026 | 0.036 | 0.924 | 0.012 | 0.223 | 0.869 | 0.149 | 0.853 |
| ≤50 | 0.209 | 0.773 | 0.028 | 0.103 | 0.841 | 0.016 | 0.346 | 0.818 | 0.288 | 0.765 |
| ≤55 | 0.372 | 0.645 | 0.032 | 0.170 | 0.751 | 0.017 | 0.473 | 0.678 | 0.423 | 0.657 |
| ≤60 | 0.549 | 0.490 | 0.032 | 0.330 | 0.605 | 0.021 | 0.634 | 0.561 | 0.596 | 0.498 |
| ≤65 | 0.720 | 0.324 | 0.032 | 0.567 | 0.427 | 0.024 | 0.784 | 0.393 | 0.688 | 0.329 |
| ≤70 | 0.864 | 0.173 | 0.031 | 0.773 | 0.243 | 0.025 | 0.871 | 0.236 | 0.832 | 0.181 |
| ≤75 | 0.940 | 0.068 | 0.030 | 0.876 | 0.099 | 0.024 | 0.938 | 0.080 | 0.933 | 0.067 |
| ≤80 | 0.978 | 0.018 | 0.030 | 0.969 | 0.030 | 0.025 | 0.985 | 0.009 | 0.966 | 0.022 |

**SNCA/VPS35**
|  | ROPAD dataset |  |  | PD GENERation dataset |  |  | MDSGene dataset |  | GP2 dataset |  |
| --- | --- | --- | --- | --- | --- | --- | --- | --- | --- | --- |
| AAO | Sens. | Spec. | PPV | Sens. | Spec. | PPV | Sens. | Spec. | Sens. | Spec. |
| ≤30 | 0.111 | 0.987 | 0.019 | N/A | 0.996 | N/A | 0.063 | 0.994 | N/A | N/A |
| ≤35 | 0.148 | 0.970 | 0.011 | N/A | 0.986 | N/A | 0.148 | 0.980 | N/A | N/A |
| ≤40 | 0.259 | 0.935 | 0.009 | 0.154 | 0.966 | 0.007 | 0.250 | 0.937 | N/A | N/A |
| ≤45 | 0.296 | 0.870 | 0.005 | 0.385 | 0.924 | 0.008 | 0.391 | 0.869 | N/A | N/A |
| ≤50 | 0.444 | 0.773 | 0.004 | 0.538 | 0.859 | 0.006 | 0.563 | 0.818 | N/A | N/A |
| ≤55 | 0.704 | 0.645 | 0.004 | 0.769 | 0.751 | 0.005 | 0.672 | 0.678 | N/A | N/A |
| ≤60 | 0.741 | 0.490 | 0.003 | 0.769 | 0.605 | 0.003 | 0.758 | 0.561 | N/A | N/A |
| ≤65 | 0.889 | 0.324 | 0.003 | 0.923 | 0.427 | 0.003 | 0.836 | 0.393 | N/A | N/A |
| ≤70 | 1.000 | 0.173 | 0.003 | 0.923 | 0.243 | 0.002 | 0.930 | 0.236 | N/A | N/A |
| ≤75 | 1.000 | 0.068 | 0.002 | 0.923 | 0.099 | 0.002 | 0.969 | 0.080 | N/A | N/A |
| **≤80** | 1.000 | 0.018 | 0.002 | 0.923 | 0.030 | 0.002 | 0.992 | 0.009 | N/A | N/A |
| **GBA1** |  |  |  |  |  |  |  |  |  |  |
|  | **ROPAD dataset** |  |  | **PD GENERation dataset** |  |  | **MDSGene dataset** |  | **GP2 dataset** |  |
| **AAO** | Sens. | Spec. | PPV | Sens. | Spec. | PPV | Sens. | Spec. | Sens. | Spec. |
| **≤30** | 0.015 | 0.987 | 0.128 | 0.005 | 0.996 | 0.104 | 0.034 | 0.994 | 0.010 | 0.983 |
| **≤35** | 0.031 | 0.970 | 0.109 | 0.013 | 0.986 | 0.074 | 0.080 | 0.980 | 0.037 | 0.958 |
| **≤40** | 0.090 | 0.935 | 0.141 | 0.050 | 0.966 | 0.111 | 0.172 | 0.937 | 0.082 | 0.912 |
| **≤45** | 0.177 | 0.870 | 0.140 | 0.110 | 0.924 | 0.110 | 0.298 | 0.869 | 0.163 | 0.853 |
| **≤50** | 0.298 | 0.773 | 0.136 | 0.206 | 0.859 | 0.111 | 0.451 | 0.818 | 0.282 | 0.765 |
| **≤55** | 0.453 | 0.645 | 0.132 | 0.329 | 0.751 | 0.101 | 0.575 | 0.678 | 0.416 | 0.657 |
| **≤60** | 0.620 | 0.490 | 0.127 | 0.531 | 0.605 | 0.103 | 0.732 | 0.561 | 0.571 | 0.498 |
| **≤65** | 0.783 | 0.324 | 0.122 | 0.689 | 0.427 | 0.093 | 0.856 | 0.393 | 0.717 | 0.329 |
| **≤70** | 0.892 | 0.173 | 0.114 | 0.835 | 0.243 | 0.086 | 0.929 | 0.236 | 0.835 | 0.181 |
| **≤75** | 0.961 | 0.068 | 0.110 | 0.940 | 0.099 | 0.082 | 0.975 | 0.080 | 0.935 | 0.067 |
| **≤80** | 0.989 | 0.018 | 0.108 | 0.981 | 0.030 | 0.079 | 0.988 | 0.009 | 0.983 | 0.022 |

## Discussion

This study assessed the diagnostic accuracy of the current selection criteria for genetic testing of the major genetic forms of PD, based on early AAO and FH, in over 25,000 patients from four large PD datasets. Across datasets, only a minority of variant carriers—especially LRRK2 and GBA1, the principal targets of gene-based trials—are captured by current recommendations.

Current genetic testing recommendations focus on patients with an AAO ≤50 years, yet this threshold only captures 23–32% of all patients with a pathogenic variant in the “real-world” datasets from the PD GENEration and ROPAD study. Accordingly, most mutation carriers—between 68% and 77%—present with later onset, and would therefore fall outside of current testing indications. Even if *GBA1* is disregarded, only 27%-38% of all monogenic PD patients are captured.

The sensitivity varied substantially between genes, capturing 79-85% of patients with recessive forms, 21-30% for *GBA1,* and only 10-21% of *LRRK2* pathogenic variant carriers (**Table 3**). This is salient because most ongoing and upcoming gene-specific clinical trials are focused on *GBA1* and *LRRK2*, the genes with the lowest variant detection rates under current testing criteria. This disconnect between research and clinical implementation may limit access to gene-specific trials and therapies.

In contrast, the MDSGene literature-based dataset yields a much higher sensitivity of 63%, nearly double that of the real-world datasets. This is most likely due to an overrepresentation of recessive forms in this dataset (≍45% *vs* ≍1% in the ROPAD and PDGENEreation datasets, **Table 1**), for which early AAO-based selection performs well. In addition, a strong publication bias toward younger-onset cases lowers the reported AAO across all genetic subtypes – including *LRRK2* – further inflating sensitivity estimates and limiting the dataset’s generalizability (**Table 1**).

Current recommendations may overprioritize early-onset forms and overlook the true age spectrum of other, more frequent genetic forms of PD in clinical reality. Specifically, late-onset genetic PD patients remain underrepresented in published case reports and series, leading to skewed assumptions about the typical AAO and potentially influencing testing guidelines. PRKN-PD illustrates this bias: only about one-seventh of reported cases had juvenile onset, yet nearly half of experts classified it as such.^16^. Thus, the MDSGene dataset, summarizing the literature, is biased toward younger-onset and recessive PD, limiting generalizability. In contrast, based on prospective genetic screening, the ROPAD and PD GENEration studies more accurately reflect the wider spectrum of AAO of genetic PD in clinical practice and are more representative of real-world testing scenarios, though these cohorts may be moderately enriched in genetic forms of PD.

Including information about FH slightly improves the AUC in all datasets, except for those involving recessive genes (**Table 2**, **Figure 1**). However, in the absence of a known pathogenic variant or given a strong *a priori* suspicion for a specific gene, there is almost no difference between AUCs when patients are preselected for having a positive FH (**Table 2**). This limited added value was expected for recessive genes, but it seems counterintuitive for dominant genes. The limited added value of FH for *LRRK2* and *GBA1* carriers is likely explained by age-related penetrance. Because AAO and FH are correlated, much of FH’s predictive value is redundant. The high prevalence of reported family history even in IPD, the typically late onset in *LRRK2*-PD, and the reduced penetrance of many dominant variants reduce the discriminatory power of FH in general testing scenarios. Variations in the definition of FH—whether restricted to first-degree relatives or not—had minimal impact on diagnostic performance metrics. Notably, classification performance based on AAO and FH may be slightly overestimated, as the weights in the logistic regression model were derived from the same datasets and were not externally validated.

PPV yielded high values for testing at AAO ≤30 (33% ROPAD; 50% PD GENEration), which declined when the threshold was raised to AAO ≤50 (20% ROPAD; 17% PD GENEration). However, these values still indicate a positive return of around 1 in 5 genetic tests for the latter threshold. Raising the threshold to AAO ≤60 would decrease the PPV by only 2%. If every PD patient were tested, the PPV would equal the prevalence of genetic PD of around 15%.^1,2^

Younger patients disproportionately drive these PPV averages, which may mislead clinicians about individual diagnostic yield, especially in older patients. For clinicians, the question is not “Whom did we capture among all younger patients?” but rather “What is *my patient’s* likelihood of carrying a pathogenic variant?”. The presented agelZlgroup stratification shows that, even in older age brackets such as an AAO between 60-70, the PPV remains substantial at 10%, which we consider clinically meaningful (**eTable 1**). Older age alone should therefore not exclude patients from testing, particularly when identifying candidates for gene-specific therapies. While AAO remains a moderate predictor, a broader AAO strategy may better align testing practices with emerging clinical trial and, eventually, therapeutic opportunities. Where broader testing is not feasible, stratification using additional biomarkers—such as α-synuclein aggregation assays or olfactory function—may help guide selection.

Equitable access to testing remains a major global challenge. Many regions – particularly across Africa, Asia, and parts of Latin America – lack access to clinical trials, and genetic testing may not be readily available or require out-of-pocket payment. These structural barriers risk further widening existing care disparities. Most ongoing gene-specific clinical trials also exclude these regions, limiting participation even among eligible carriers. Global harmonization of access to genetic diagnostics and precision therapies should be prioritized alongside developing clinical testing guidelines, which is one of the aims of GP2 and the MDS.

A key strength of this study lies in its unprecedented scale, encompassing data from over 25,000 individuals with PD. This enables the most comprehensive analysis of genetic testing approaches to date.

Several limitations should be acknowledged. Four distinct datasets allow differentiation between literature, global cohort-based, and ‘real-world’ data, considering different definitions of FH. Ancestry, which can influence the probability of carrying a pathogenic PD variant, was not considered. While the ROPAD and PD GENEration studies most closely reflect real-world conditions, they still are research-based and subject to participation and selection biases. To avoid overestimating diagnostic performance, we applied a cautious approach: individuals with variants in more than one gene or genes linked to atypical PD were excluded, and variants in recently proposed PD genes such as *RAB32* and other novel candidates were not included, as they were not uniformly analyzed across all datasets.^21^

The genetic screening process differed across datasets, mainly relying on panel analyses or genotyping rather than whole-genome sequencing, which limited the detection of certain variant types. Copy-number variants were not uniformly assessed and were absent from the GP2 dataset, potentially contributing to an underestimation of genetic PD and misclassification of some carriers as IPD. Nevertheless, this conservative design enhances real-world applicability by reflecting current diagnostic limitations. Finally, as most individuals in the study were of European ancestry, further studies will be necessary to determine how our conclusions generalize to more genetically diverse populations.

In conclusion, current criteria for genetic testing, such as testing every individual with an AAO ≤50 years, fail to identify a substantial proportion of patients with pathogenic variants. This study provides data-driven estimates of sensitivity and PPV at various age thresholds, which can serve as the foundation for developing data-driven recommendations for genetic testing in PD. While PPVs are not a direct measure of clinical or economic effectiveness, they provide important context for the diagnostic yield and resource implications of broader testing efforts – particularly in the context of gene-targeted trials. As such therapies become available, genetic testing will likely be offered to all PD patients wherever possible, as already seen in other neurological diseases.^22^

## Data Availability

Data used in the preparation of this article were obtained, among others, from the Global Parkinson's Genetics Program (GP2; https://gp2.org). Specifically, we used Tier 2 data from GP2 release 9 (DOI 10.5281/zenodo.14510099). GP2 data are available on AMP PD (https://amp-pd.org). Data were also obtained from ROPAD, PD GENEration, the MDSGene database, and two German observational studies (EPIPARK and DeNoPa). These datasets can be accessed by individual request to the corresponding author.
All code generated for this article, and the identifiers for all software programs and packages used, are available on GitHub (https://github.com/GP2code/Genetic_Testing_in_PD) and were given a persistent identifier via Zenodo (DOI: 10.5281/zenodo.17567723).

## Data and code availability

Data used in the preparation of this article were obtained, among others, from the Global Parkinson’s Genetics Program (GP2; https://gp2.org). Specifically, we used Tier 2 data from GP2 release 9 (DOI 10.5281/zenodo.14510099). GP2 data are available on AMP PD (https://amp-pd.org). Data were also obtained from ROPAD, PD GENEration, the MDSGene database, and two German observational studies, these datasets can be accessed by individual request to the corresponding author.

All code generated for this article, and the identifiers for all software programs and packages used, are available on GitHub (https://github.com/GP2code/Genetic_Testing_in_PD) and were given a persistent identifier via Zenodo (DOI: 10.5281/zenodo.17567723).

## Acknowledgement

This project was supported by the Global Parkinson’s Genetics Program (GP2; https://gp2.org). GP2 is funded by the Aligning Science Across Parkinson’s (ASAP) initiative and implemented by The Michael J. Fox Foundation for Parkinson’s Research (MJFF). For a complete list of GP2 members, see doi.org/10.5281/zenodo.7904831.

This research was partly supported by the Intramural Research Program of the National Institutes of Health (NIH). The contributions of the NIH author(s) were made as part of their official duties as NIH federal employees, are in compliance with agency policy requirements, and are considered Works of the United States Government. However, the findings and conclusions presented in this paper are those of the author(s) and do not necessarily reflect the views of the NIH or the U.S. Department of Health and Human Services.

We thank the MDSGene curators for their continuous efforts in compiling and annotating genetic and clinical data from the literature.

For the purpose of Open Access, the author has applied a CC BY public copyright license to any Author Accepted Manuscript version arising from this submission.

