## Supplementary Methods for "How many do we miss? - Evaluation of age at onset and family history as selection criteria for genetic testing in Parkinson’s disease"

**eMethods 1: The ROPAD Study Dataset**

The ROPAD study (Rostock Parkinson’s Disease) was a prospective, multicenter genetic screening study conducted across 16 countries.^1,2^ A total of 12,580 individuals with a clinical diagnosis of Parkinson’s disease (PD) were enrolled. Genetic testing was performed by CENTOGENE GmbH, a certified diagnostic laboratory, following a standardized, three-tiered protocol:

Tier 1: *LRRK2* and *GBA1* screening

All participants were initially screened for 11 predefined pathogenic or likely pathogenic variants in the *LRRK2* gene:

p.Gly2019Ser, p.Arg1441Cys, p.Arg1441Gly, p.Arg1441His, p.Ile2020Thr, p.Tyr1699Cys, p.Asn1437His, p.Ile1371Val, p.Leu1165Pro, p.Glu1874Ter, and p.Gly2385Arg.
This was followed by comprehensive sequencing of *GBA1*, using long-range PCR and exon-specific PCR combined with next-generation sequencing (NGS). This approach enabled the detection of single-nucleotide variants and complex alleles, including recombinant alleles. Participants with negative findings in Tier 1 proceeded to the next stage.

Tier 2: 68-Gene Panel Sequencing

Participants negative for *LRRK2* and *GBA1* variants underwent NGS-based sequencing of a predefined 68-gene panel. This panel included genes with established or suspected involvement in PD or related neurodegenerative conditions. The following genes were included: *ACMSD, ADH1C, APOE, ATP13A2, ATP1A3, CHCHD2, C19orf12, COASY, CYP2D6, DNAJC6, DNMT1, EIF4G1, FBXO7, GBA, GCH1, GRN, HTRA2, LRRK2, MAPT, MCCC1, MIR137, MIR4697, MTHFR, NEU1, NR4A2, NPC1, PANK2, PRKN), PARK7 (DJ1), PINK1, PLA2G6, POLG, PRKRA, PSEN1, PVRL3, RAB39B, SLC6A3, SLC20A2, SLC30A10, SLC39A14, SLC41A1, SNCA, SPG11, SPG15, SPG20, SPG21, SPG35, SYNJ1, TAF1, TH, TMEM230, TMEM240, TREM2, TSEN54, TUBB4A, UBQLN2, UCHL1, USP24, VPS13C, VPS35, WDR45, XK, ZFYVE26, ZNHIT3, ZNRF3, DNAJC13, HLA-DRB5, GCHFR, HTR2A.*

Panel sequencing was performed using hybridization-based target enrichment and Illumina sequencing platforms. Copy number variant (CNV) detection was performed using a validated, clinically implemented internal CNV-calling algorithm. Participants with negative findings in Tier 2 proceeded to the next stage.

Tier 3: Whole Genome Sequencing (WGS)

Participants with no reportable variants in Tiers 1 or 2 and with high clinical suspicion for monogenic PD—defined by age at onset <56 years and/or positive family history—were offered short-read WGS after providing additional consent. WGS was performed at ≥30× coverage with variant calling for single-nucleotide variants (SNVs), indels, and structural variants.

Clinical Data Collection and Reporting

Age at onset (AAO) was defined as the age at which the first cardinal motor symptom of PD appeared, as reported by the patient or documented by the treating physician. Family history (FH) was considered positive if any biologically related family member had been diagnosed with PD, regardless of generational distance. Clinical data were documented via standardized electronic case report forms. Variant interpretation followed ACMG-AMP guidelines. Reportable variants (classified as pathogenic, likely pathogenic, or risk variants) were returned to participants, and post-test genetic counseling was provided in all cases.

**eMethods 2: The PD GENEration Study Dataset**

The PD GENEration study^3^ is an ongoing, prospective, multi-center observational initiative led by the Parkinson’s Foundation (NCT04994015) and supported by GP2 and the Aligning Science Across Parkinson’s (ASAP) initiative implemented by the Michael J. Fox Foundation for Parkinson’s Research. It offers free-of-charge, diagnostic-grade genetic testing and genetic counseling to individuals with a clinical diagnosis of Parkinson’s disease (PD) across the United States, Canada, and the Dominican Republic. Participants are enrolled either through in-person visits at participating clinical centers or remotely through a secure digital platform.

Genetic testing is performed by Fulgent Genetics, a CLIA- and CAP-certified laboratory, using next-generation sequencing (NGS) with full exon coverage and deletion/duplication (CNV) analysis. The panel includes the following seven genes with established pathogenic roles or significant risk associations in PD:*GBA1, LRRK2, PRKN (PARK2), PINK1, PARK7 (DJ1), SNCA, and VPS35*.
All coding regions and exon–intron boundaries of these genes are analyzed. Structural variants and large deletions/duplications are detected using a combination of read-depth and split-read algorithms validated for clinical diagnostics.

Participants receive pre- and post-test genetic counseling, and all results meeting clinical reporting thresholds (i.e., pathogenic, likely pathogenic, or risk variants) are returned. Variant interpretation is performed according to ACMG-AMP criteria, and classification includes explicit annotation of risk-associated alleles (e.g., *GBA1* p.N370S, *LRRK2* p.G2019S).

Clinical data include demographic details, age at onset (AAO), family history (FH), and prior diagnostic history. AAO is defined as the age at which the first motor symptom attributable to PD occurred. Family history is defined as having at least one first-degree relative diagnosed with PD.

The PD GENEration dataset was directly shared with authors by the PD GENEration study team and represented data compiled up to May 1, 2023. PD GENEration data is separately available via the AMP-PD data portal (amp-pd.org).

**eMethods 3: MDSGene Study Dataset**

Genetic PD patients: MDSGene

The MDSGene study dataset was derived from the International Parkinson’s Disease and Movement Disorder Society Genetic Mutation Database (mdsgene.org), a manually curated, literature-based repository of genetically confirmed PD patients. Only individuals reported to carry potentially disease causing variants pathogenic in monogenic PD genes (*PRKN*^4^*, PINK1*^4^*, PARK7/DJ-1*^4^*, LRRK2*^5^*, SNCA*^5,6^*, VPS35*^5^) or *GBA1*^7^ were included.

Potentially disease-causing variants are defined as definitively and probably pathogenic after the MDSGene criteria. Each entry was curated from peer-reviewed literature and required individual-level clinical annotation, including AAO and FH. No overlap exists between MDSGene and the ROPAD or PD GENEration cohorts, since these cohorts were published after the last MDSGene update. However, due to the publicly curated nature of MDSGene, overlap with GP2 data cannot be completely excluded.

IPD patients: EPIPARK and DeNoPa cohort studies

EPIPARK was a population-based observational cohort study established to investigate the prevalence, development, and impact of non-motor symptoms in Parkinsonism, including Parkinson’s disease (PD), in Northern Germany. The study was initiated in Lübeck, Germany, and aimed to characterize a representative sample of individuals newly diagnosed with PD, capturing both clinical and epidemiological data longitudinally. Details of the study protocol have been published previously^8^.

Eligible participants were required to have a clinical diagnosis of idiopathic PD based on UK Brain Bank criteria, be within 12 months of diagnosis, and not have undergone dopaminergic treatment for longer than 6 months. Recruitment began in 2011, and participants underwent structured assessments including comprehensive interviews, neurological examinations, and standardized questionnaires focused on non-motor symptoms, motor phenotype, and quality of life. Genetic testing was performed using a gene panel targeting known PD genes (similar to the panel used for the ROPAD study, including *LRRK2, PRKN, PINK1, PARK7/DJ1, SNCA, VPS35*, and *GBA1*). Only patients in whom no pathogenic or likely pathogenic variants in these genes were identified were included as idiopathic PD (IPD) controls for the MDS Gene study dataset.

The De Novo Parkinson (DeNoPa) cohort was a prospective, single-center observational study of newly diagnosed, drug-naïve PD patients. The study was designed to explore clinical progression and biomarkers of early PD. Recruitment began in 2009 at the Paracelsus-Elena Clinic in Kassel, Germany, and the study design and initial findings have been reported elsewhere^9^.

Participants were enrolled within 6 months of PD diagnosis and had not received dopaminergic medication prior to baseline assessment. The diagnosis of PD was confirmed according to UK Brain Bank criteria and supported by neuroimaging (e.g., DaTSCAN). The study included longitudinal follow-up with standardized evaluations of motor and non-motor symptoms, neuropsychological performance, and biosample collection (blood, CSF). DeNoPa participants underwent the same genetic testing as in EPIPARK. Only those patients without pathogenic or likely pathogenic variants were classified as idiopathic PD (IPD) and included in the MDSGene dataset.

**eMethods 4:** **GP2 Study Dataset**

The Global Parkinson’s Genetics Program (GP2; <https://gp2.org>) is an international collaborative initiative aimed at better understanding the genetic architecture of PD and parkinsonism by performing genome-wide genotyping and short-read genome sequencing at a large scale. GP2’s overall workflow, including site recruitment and data and sample collection, has been described before^10–12^.

For this project, we used genome-wide genotyping data from GP2’s Release 9 (DOI 10.5281/zenodo.14510099). The release incorporated genotyping data of a total of 68,249 individuals from eleven different genetically determined ancestries, including affected individuals with PD, atypical parkinsonism and other neurodegenerative phenotypes, and unaffected individuals (healthy controls, unaffected family members, and population cohorts). Genotyping was performed using the Illumina NeuroBooster Array (NBA).^13^ Raw genotyping data underwent quality control and genetic ancestry prediction using GenoTools v1.2.3 with the default settings.^12^ Variants in the genes of interest were extracted using PLINK^14,15^ and annotated with ANNOVAR.^16,17^ Genes of interest included *LRRK2, SNCA, VPS35, PINK1, PRKN,* and *PARK7/DJ1*, all linked to monogenic forms of PD, as well as *GBA1* as the strongest PD risk gene. We only included variants evaluated to be pathogenic or likely pathogenic according to ClinVar (<https://www.ncbi.nlm.nih.gov/clinvar/>)^18^ and/or the consensus recommendations of the American College of Medical Genetics and Genomics (ACMG)^19^. Data submitted to GP2 from ROPAD and PD GENEration were explicitly excluded by removing these from the dataset; overlap with MDSGene cannot be ruled out, as there is the possibility that individual patients have been published and sent for inclusion in the GP2 dataset.

Patients that were not classified as PD were excluded from the dataset, as well as carriers of variants in two or more different genes. The GP2 data analysis did not include CNV analysis.

**Supplementary Tables**

**eTable 1. Positive Predictive Value, Sensitivity and Specificity of Genetic Testing at Different Age of Onset Timeframes (A) ROPAD Dataset (B) PD GENEration Dataset.**

AAO = age at onset; PPV = positive predictive value

**A**

| **AAO (years)** | **Total tested (n)** | **Genetic positive (n)** | **Genetic positive without *GBA 1* (n)** | ***PARK7***  **(n)** | ***GBA1***  **(n)** | ***LRRK2***  **(n)** | ***PINK1***  **(n)** | ***PRKN***  **(n)** | ***SNCA***  **(n)** | ***VPS35***  **(n)** | **PPV** | **Sensitivity** | **Specificity** | **PPV**  **without *GBA1*** | **Sensitivity**  **without *GBA1*** | **Specificity**  **without *GBA1*** |
| --- | --- | --- | --- | --- | --- | --- | --- | --- | --- | --- | --- | --- | --- | --- | --- | --- |
| **<30** | 193 | 63 | 41 | 2 | 20 | 2 | 3 | 33 | 3 | 0 | 0.326 | 0.034 | 0.987 | 0.212 | 0.079 | 0.96 |
| **30–40** | 713 | 164 | 65 | 2 | 97 | 16 | 4 | 41 | 4 | 0 | 0.230 | 0.090 | 0.947 | 0.091 | 0.125 | 0.96 |
| **40–50** | 2,036 | 363 | 91 | 0 | 272 | 59 | 2 | 25 | 5 | 0 | 0.178 | 0.198 | 0.839 | 0.045 | 0.174 | 0.958 |
| **50–60** | 3,508 | 570 | 149 | 0 | 421 | 125 | 0 | 16 | 7 | 1 | 0.162 | 0.311 | 0.717 | 0.042 | 0.285 | 0.957 |
| **60–70** | 3,775 | 480 | 125 | 0 | 355 | 116 | 0 | 2 | 6 | 1 | 0.127 | 0.262 | 0.682 | 0.033 | 0.239 | 0.953 |
| **70–80** | 1,780 | 170 | 43 | 0 | 127 | 42 | 0 | 1 | 0 | 0 | 0.096 | 0.093 | 0.845 | 0.024 | 0.082 | 0.954 |
| **>80** | 204 | 22 | 8 | 0 | 14 | 8 | 0 | 0 | 0 | 0 | 0.108 | 0.012 | 0.982 | 0.039 | 0.015 | 0.957 |

**B**

| **AAO (years)** | **Total tested (n)** | **Genetic positive (n)** | **Genetic positive without *GBA 1* (n)** | ***PARK7* (n)** | ***GBA1* (n)** | ***LRRK2* (n)** | ***PINK1* (n)** | ***PRKN* (n)** | ***SNCA* (n)** | ***VPS35* (n)** | **PPV** | **Sensitivity** | **Specificity** | **PPV**  **without *GBA1*** | **Sensitivity**  **without *GBA1*** | **Specificity**  **without *GBA1*** |
| --- | --- | --- | --- | --- | --- | --- | --- | --- | --- | --- | --- | --- | --- | --- | --- | --- |
| **<30** | 50 | 25 | 22 | 2 | 3 | 0 | 1 | 19 | 0 | 0 | 0.500 | 0.028 | 0.890 | 0.468 | 0.082 | 0.996 |
| **30–40** | 262 | 47 | 19 | 0 | 28 | 2 | 0 | 15 | 2 | 0 | 0.179 | 0.053 | 0.890 | 0.081 | 0.071 | 0.969 |
| **40–50** | 879 | 129 | 32 | 0 | 97 | 15 | 3 | 9 | 4 | 1 | 0.147 | 0.145 | 0.892 | 0.041 | 0.119 | 0.893 |
| **50–60** | 2037 | 259 | 58 | 0 | 201 | 47 | 0 | 8 | 1 | 2 | 0.127 | 0.291 | 0.892 | 0.032 | 0.216 | 0.746 |
| **60–70** | 2812 | 280 | 91 | 0 | 189 | 86 | 0 | 3 | 1 | 1 | 0.100 | 0.315 | 0.880 | 0.035 | 0.338 | 0.638 |
| **70–80** | 1620 | 130 | 40 | 0 | 90 | 38 | 0 | 2 | 0 | 0 | 0.080 | 0.146 | 0.879 | 0.026 | 0.149 | 0.787 |
| **>80** | 232 | 19 | 7 | 0 | 12 | 6 | 0 | 0 | 1 | 0 | 0.082 | 0.021 | 0.886 | 0.032 | 0.026 | 0.97 |

**eTable 2. Statistical Comparison of ROC Curve Performance Based on Age at Onset Alone vs. Combined with Family History Across Four Cohorts.**

AAO = age at onset; FH = family history; AUC = area under the curve; ΔAUC = delta between both area under the curves; CI = confidence interval; PR AUC = precision recall AUC; N/A = no calculations were performed due to the small n

| **Dataset** | **Group** | **AUC**  **(AAO)** | **AUC**  **(AAO + FH)** | **ΔAUC** | **95% CI of ΔAUC** | **DeLong p** | **PR AUC** | **Carrier prevalence** |
| --- | --- | --- | --- | --- | --- | --- | --- | --- |
| **ROPAD** | All genetic | 0.590 | 0.600 | 0.015 | [-0.023, -0.006] | <0.001 | 0.200 | 0.150 |
|  | *PRKN/PINK1/PARK7* | 0.902 | 0.901 | -0.001 | [-0.005, 0.006] | 0.805 | 0.148 | 0.010 |
|  | *GBA1* | 0.574 | 0.579 | 0.005 | [-0.013, 0.003] | 0.234 | 0.134 | 0.107 |
|  | *LRRK2* | 0.521 | 0.606 | 0.084 | [-0.114, -0.055] | <0.001 | 0.034 | 0.030 |
|  | *VPS35/SNCA* | 0.709 | 0.817 | 0.108 | [-0.197, -0.019] | 0.017 | 0.007 | 0.002 |
|  | All genes without *GBA 1* | 0.623 | 0.673 | 0.050 | [-0.062, -0.026] | <0.001 | 0.102 | 0.043 |
| **PD GENEration** | All genetic | 0.581 | 0.600 | 0.019 | [-0.032, -0.006] | 0.005 | 0.169 | 0.113 |
|  | *PRKN/PINK1/PARK7* | 0.893 | 0.903 | 0.010 | [-0.024, 0.004] | 0.165 | 0.358 | 0.008 |
|  | *GBA1* | 0.580 | 0.582 | 0.002 | [-0.011, 0.006] | 0.583 | 0.101 | 0.079 |
|  | *LRRK2* | 0.471 | 0.639 | 0.168 | [-0.236, -0.100] | <0.001 | 0.024 | 0.025 |
|  | *VPS35/SNCA* | 0.782 | 0.835 | 0.053 | [-0.145, 0.040] | 0.269 | 0.007 | 0.002 |
|  | All genes without *GBA 1* | 0.584 | 0.657 | 0.073 | [-0.102, -0.045] | <0.001 | 0.126 | 0.034 |
| **MDSGene** | All genetic | 0.789 | 0.831 | 0.043 | [-0.058, -0.027] | <0.001 | 0.961 | 0.878 |
|  | *PRKN/PINK1/PARK7* | 0.945 | 0.952 | 0.007 | [–0.012, –0.002] | 0.007 | 0.980 | 0.362 |
|  | *GBA1* | 0.686 | 0.72 | 0.034 | [–0.060, –0.007] | 0.013 | 0.761 | 0.195 |
|  | *LRRK2* | 0.640 | 0.748 | 0.108 | [–0.142, –0.075] | <0.001 | 0.769 | 0.277 |
|  | *VPS35/SNCA* | 0.749 | 0.866 | 0.117 | [–0.159, –0.075] | <0.001 | 0.515 | 0.044 |
|  | All genes without *GBA 1* | 0.802 | 0.852 | 0.05 | [–0.060, –0.029] | <0.001 | 0.958 | 0.684 |
| **GP2** | All genetic | 0.552 | 0.578 | 0.026 | [-0.044, -0.008] | 0.005 | 0.243 | 0.224 |
|  | *PRKN/PINK1/PARK7* | 0.881 | 0.905 | 0.024 | [–0.067, 0.019] | 0.28 | 0.044 | 0.005 |
|  | *GBA1* | 0.546 | 0.545 | 0 | [–0.015, 0.016] | 0.93 | 0.194 | 0.173 |
|  | *LRRK2* | 0.537 | 0.663 | 0.127 | [–0.168,  –0.086] | <0.001 | 0.057 | 0.045 |
|  | *VPS35/SNCA* | N/A | N/A | N/A | N/A | N/A | N/A | N/A |
|  | All genes without *GBA 1* | 0.567 | 0.687 | 0.121 | [–0.152, –0.076] | <0.001 | 0.075 | 0.051 |

**Supplementary Figures**

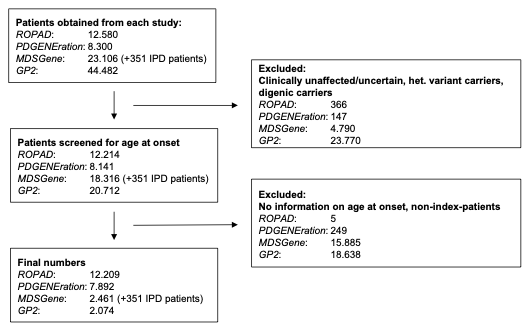

**eFigure 1. Flowchart of the exclusion of patients for the four individual datasets.**

IPD = Idiopathic Parkinson’s disease
